# Rapid Online Assessment of Reading (ROAR): Evaluation of an Online Tool for Screening Reading Skills in a Developmental-Behavioral Pediatrics Clinic

**DOI:** 10.1101/2023.05.10.23289794

**Authors:** Elizabeth P. Barrington, Sadie Mae Sarkisian, Heidi M. Feldman, Jason D. Yeatman

## Abstract

**Objective:** Reading difficulties frequently co-occur with other neurodevelopmental/behavioral conditions. It is difficult to assess reading routinely in Developmental-Behavioral Pediatrics (DBP) clinical practice due to time/resource constraints. Rapid Online Assessment of Reading (ROAR) is a gamified assessment that children take in a web-browser without adult supervision. This study’s purpose was to evaluate ROAR as a screening tool for reading difficulties in a DBP clinic.

**Method:** Patients, ages 6-14 years, attending a DBP clinic, were invited to participate. Children took ROAR and completed the Woodcock-Johnson Letter-Word Identification (LWID) and Word Attack (WA). Basic Reading Skills (BRS), a standardized aggregate score of LWID and WA, was used as the gold standard assessment. The strength of association between age-adjusted standard score on ROAR and BRS was calculated. BRS scores < 90 (bottom quartile of sample) were deemed poor reader. Receiver Operating Characteristic (ROC) curve analysis was used to assess the quality of ROAR as a screening test.

**Results:** A total of 41 children, 78% boys, mean age 9.5 years (SD 2.0 years), completed the study. The correlation of ROAR standard score with BRS was r = 0.66, p<0.001. ROC curve classification analysis with ROAR scores accurately classified poor readers with an area under the curve (AUC) of 0.90.

**Conclusion:** ROAR is a useful screening tool for children to take before attending a DBP clinic to identify children at high risk for reading difficulties. Assessment of the tool during a busy clinic was challenging and a larger replication is warranted.

## BACKGROUND

Reading disorders (e.g., dyslexia) affect 5-12% of the general population^1–3^. Children with reading disorders are at increased risk for difficulties across academic domains, including writing and math, and for broader struggles in school generally^1,2^. Reading disorders often co-occur with other neurodevelopmental and neurobehavioral conditions. For example, the prevalence of reading disorders is estimated to be 25-40% in children with attention deficit-hyperactivity disorder (ADHD)^4,5^, approximately 30% in children with autism^6^ and 10% in children with anxiety^7^. Developmental-Behavioral Pediatrics (DBP) clinics frequently evaluate children with ADHD, autism, anxiety, and related neurobehavioral conditions. Because of the high prevalence of co-occurrence, evaluation of reading skills during a DBP evaluation would be useful for developing a thorough understanding of a child’s learning profile and for designing a comprehensive management plan. However, it is difficult to routinely include a reading assessment in busy clinical practice due to time constraints and lack of resources. A valid screening tool that accurately identifies children at high risk for reading disorders would be useful in DBP clinical practice for targeting which children should have a reading assessment, particularly if the screening results could be available to clinicians at the time of the initial evaluation.

The Rapid Online Assessment of Reading Screening (ROAR)^8^ is an online reading screening tool, developed for the general population of school-aged children. The tool uses a two-alternative, forced choice, time-limited lexical decision task (LDT) to assess reading skills^8^. The task has been gamified to be engaging and easy to navigate for school-aged children and adolescents. The child is automatically led through a series of increasingly difficult printed words or pronounceable non-words (pseudowords) that appear on the computer screen. The task is to indicate if the stimulus is a valid word versus a non-word, or, in the game language, a “magical word”. The test has been formatted to be delivered in a web-browser, requiring no adult involvement once the child has been enrolled and has entered the site. Thus, ROAR can be self-administered online from any computer without the presence of a parent, teacher, psychologist, or other adult. Because it is delivered through the web-browser, it does not require any specialized software to be installed. The results are relayed to a site where age-adjusted scoring occurs, and results are generated. These results can be made available for individuals or for groups. ROAR has proven to be an accurate approximation of standard reading measures in children between ages 6 and 18 years with reading profiles ranging from severely impaired to exceptional^8^. In the initial validation study, ROAR performance had high correlation (r = 0.91) with raw scores on Woodcock-Johnson Letter Word Identification (LWID), a standardized measures of reading ability^8^. ROAR has also proven to be highly reliable (r = 0.97) across test administrations^8^.

The purpose of this study was to evaluate ROAR for use in a clinical population within a DBP clinic with children that are likely to have a wide range of academic abilities and co-existing conditions that affect school functioning. We evaluated ROAR in this clinical population specifically because such children may have difficulties performing well on ROAR because of inattention, assessment anxiety, and/or other challenges common to the neurodivergent population. Because ROAR is gamified, simple, and short, we were hopeful it would prove reliable and valid in the DBP clinical population. We hypothesized that age-adjusted scores on the ROAR would be strongly correlated with age-adjusted standard scores on the gold-standard test--the Basic Reading Skills (BRS) aggregate score from the Woodcock-Johnson Tests of Achievement. The BRS combines scores from the Letter-Word Identification Test (LWID) and Word Attack (WA) to assess single word reading and phonological decoding^9^. If ROAR can be shown to correlate with established standardized reading assessments in a clinical population, then it may prove to be a useful pre-clinical screening tool that could be completed at home to accurately identify those children referred to DBP clinical care who are at high risk of concurrent reading disorders.

## METHODS

### Study Design

The study is a prospective cohort design comparing ROAR as a screening test versus a gold standard measure of reading ability.

### Participants

Participants (N=41) were recruited between August 2021 and August 2022 in the DBP clinic at [name withheld], part of the [name withheld] health care system in [city/state withheld]. Participants were patients being evaluated in the clinic for the first time and patients returning for follow-up.

### Recruitment

Potential study participants were identified by direct referral to the research team by their DBP clinician or were identified from the clinic schedule. To be eligible for the study, the caregiver needed to confirm the child could at least read simple sight words. The child also needed documented English proficiency, defined as having either English as a first language or having completed at least two years of schooling in English if a language other than English was spoken at home. Patients and families that were interested in participation in the study were given the invitation flyer that contained an internet link to an online dashboard where the parent provided written consent, HIPAA authorizations, and a brief parent questionnaire. The child provided study assent and completed ROAR. All study procedures, consent documents and recruitment materials were presented to and approved by [medical institution withheld] IRB.

### Measures

ROAR is a two-alternative, forced choice, lexical decision task delivered in a web-browser. Before beginning ROAR, the participant is led through several pages of instructions explaining how to complete the assessment with practice and feedback to ensure understanding. In addition to written instructions, each page has a voice recording of the instructions that can be heard through the computer’s speakers. This study used the same version of ROAR introduced in the original paper even though subsequent versions have been released^8^ (newer versions of ROAR use a computer adaptive testing algorithm for more efficient assessment). In this version, the instructions explain that the participant has entered the world of Lexicality and the goal is to reach a gate that connects the participant to their own world, Earth. The participant’s helper, the Scout, explains that to find the gate, the participant needs to tell the difference between the magical language of Lexicality, a set of pronounceable non-words (pseudowords), and words in English. The stimulus word flashes briefly (350ms) on the computer screen and the participant uses the arrow keys on the computer (left or right) to indicate if the word is real or magical (Figure 1a). The participant completes up to 3 practice trials, with corrections and additional practice items before the participant can continue. The participant must answer each practice trial correctly before they can advance to the actual assessment. The test consists of 252 word or pseudoword prompts split up into four sections with breaks offered in between sections. The set of prompts was determined by preliminary studies using Item Response Theory^8^. ROAR back-end programming calculates a total correct score and measures how long between word/pseudoword presentation and when a left or right arrow key was struck. When the participant answers correctly, a pleasant chime sound is produced and when the participant answers incorrectly a dissonant thud sound is produced. There is no time limit for participants to choose the right or left arrow key, but an answer is required for the game to proceed. Participants are kept engaged during ROAR by collecting coins - the participant gets 10 gold coins for each 10 correct answers (Figure 1b). In addition, for each section completed an animated character joins the participant’s journey.

**Figure 1:**
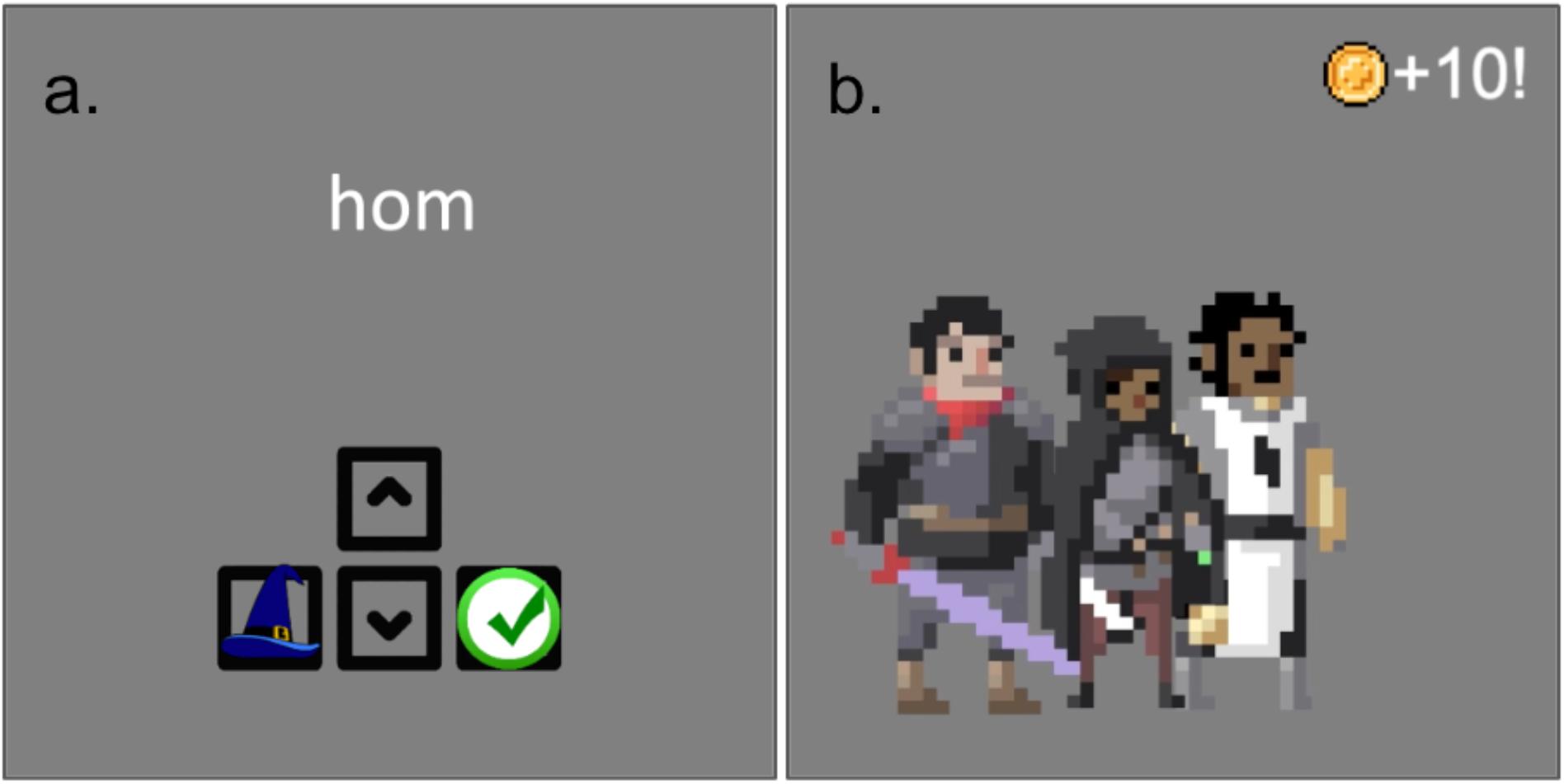
The image on the left (1a) shows an example of a pseudoword and the prompts on the screen for the participant to choose the left or right arrow key. In this case the participant would use the left arrow key to indicate that “hom” is a pseudoword. The image on the right (1b) is animated and displayed to the child after collecting 10 coins. The child is nearly at the end of ROAR testing and has three companions that have joined the adventure.

The Woodcock-Johnson Tests of Achievement Letter-Word Identification (LWID) and Word-Attack (WA)^10^ served as the gold standard assessment of single word reading ability for the study and were administered to participants by a member of the research team. Standard scores can be calculated for individual tests as well as aggregate scores. The Basic Reading Skills (BRS) score is an aggregate measure that combines scores of LWID and WA. The LWID, WA, and BRS each have a median reliability of 0.92, 0.90, and 0.95 respectively in the 5 to 18 age range^9^.

### Chart Review

The research staff performed a chart review on the 41 participants that completed the study. The information collected in chart review included: age, sex, top three diagnoses associated with the clinical encounter at time of recruitment, presence or absence of school supports such as individualized education plan (IEP) or 504 plan, and insurance type.

### Data Analysis

Descriptive statistics were tabulated to evaluate and categorize the patient population. The degree of association between the ROAR and standard scores on the LWID, WA and BRS were calculated using Pearson moment correlation. The degree of association was assessed using partial correlations with potential covariates, including the child’s sex, co-existing diagnosis, and age. To evaluate ROAR as a screening test for reading ability, we performed classification analysis with creating a receiver operating characteristic (ROC) curve and assessed area under the curve (AUC). We defined participants as poor readers if their BRS score fell in the lowest quartile of scores.

## RESULTS

Participants that completed the study were between the ages of 6 and 14 years. Table 1 summarizes characteristics of the patient cohort. The mean age of the participants was 9.5 years; the majority were male (78.6%) and had private insurance (69%). The percent who had no school support in place such as an IEP or 504 plan was 48.8%. Most participants had more than one medical diagnosis associated with the clinical encounter at which they were recruited. The four most common diagnoses were ADHD, any type (68.3%), anxiety (41.5%), learning disability (24.4%) and autism (22%). Most participants learned English as their first language (90.2%).

**Table 1:** Characteristics of Patient Cohort Aged 6-14 years (n=41)

|  |  |
| --- | --- |
| Age<sup>a</sup> (years) | Min 6.5, Max 14.2, Mean 9.5, Std Dev 2.0 |
| Male, n (%) | 33 (78.6%) |
| Insurance type, n (%) |  |
| Public | 13 (31.0%) |
| Private | 29 (69.0%) |
| Special Education Services: |  |
| No IEP/504 | 20 (48.8%) |
| IEP in place | 15 (36.6%) |
| IEP evaluation in process | 3 (7.3%) |
| 504 in place | 1 (2.4%) |
| Other/unknown | 2 (4.9%)<sup>b</sup> |
| Diagnoses per most recent medical encounter: |  |
| Single diagnosis | 14 (34.1%) |
| Two diagnoses | 18 (43.9%) |
| Three diagnoses | 7 (17.1%) |
| Symptom level diagnosis provided only | 2 (4.9%) |
| Diagnosis (total) |  |
| ADHD, any type | 28 (68.3%) |
| Anxiety | 17 (41.5%) |
| Learning Disability | 10 (24.4%) |
| Autism | 9 (22%) |
| Speech language disorder | 3 (7.3%) |
| Depression | 2 (4.9%) |
| Oppositional Defiant Disorder | 1 (2.4%) |
| Disruptive mood dysregulation disorder (DMDD) | 1 (2.4%) |
| Intellectual disability | 0 |
| Diagnoses (multiple) |  |
| ADHD and Anxiety | 9 (22.0%) |
| ADHD and Learning Disability | 6 (14.6%) |
| ADHD and Autism | 5 (12.2%) |
| ADHD and Speech Language disorder | 2 (4.9%) |
| Anxiety and Autism | 4 (9.8%) |
| Anxiety and Depression | 2 (4.9%) |
| English is primary language | 37 (90.2%)<sup>c</sup> |

<sup>a</sup>Age in years when the child completed ROAR. Mean displayed as continuous variable.
<sup>b</sup>One patient had a student study team evaluation but no IEP as of yet. Another patient's mother was unsure what if any special education resources were being provided to her child.
<sup>c</sup>Four parents did not complete the parent survey

### Correlation

ROAR raw score was strongly correlated with raw scores on selected Woodcock Johnson test of Achievement: LWID (r = 0.76, p < 0.001, CI [0.59, 0.87]) (Figure 2a) and WA (r = 0.73, p < 0.001, CI [0.55, 0.85]). There continued to be strong correlation between ROAR scores and selected Woodcock Johnson test of Achievement when ROAR scores and Woodcock Johnson scores were age-adjusted and standardized: BRS (r = 0.66, p < 0.001, CI [0.44, 0.80]) (Figure 2b), LWID (r = 0.65, p < 0.001, CI [0.42, 0.80]) and WA (r = 0.63, p < 0.001, CI [0.40, 0.79]). Scatter plots revealed three participants that were identified as outliers with a standardized residual that was larger than 2 (in absolute value). One participant was recruited during a new patient visit; this child had a poor BRS score and poor ROAR standard score and was identified as likely having intellectual disability during the clinical visit. The second outlier did well on all assessments though comparatively less well on ROAR than BRS; difficulty with sustaining attention after a long clinical visit likely contributed to lower, although still high average, ROAR score. The third outlier performed in the average range on LWID and WA in clinic then went home to complete the ROAR and performed above the average range; the parent shared that the child was somewhat inattentive in the clinic testing and at home was given a distraction free environment to work on the ROAR where he performed better than on the standardized measures. Without the three outliers the correlation between standard score on ROAR and BRS only changed slightly (r = 0.68, p < 0.001, CI [0.46, 0.82])

**Figure 2:**
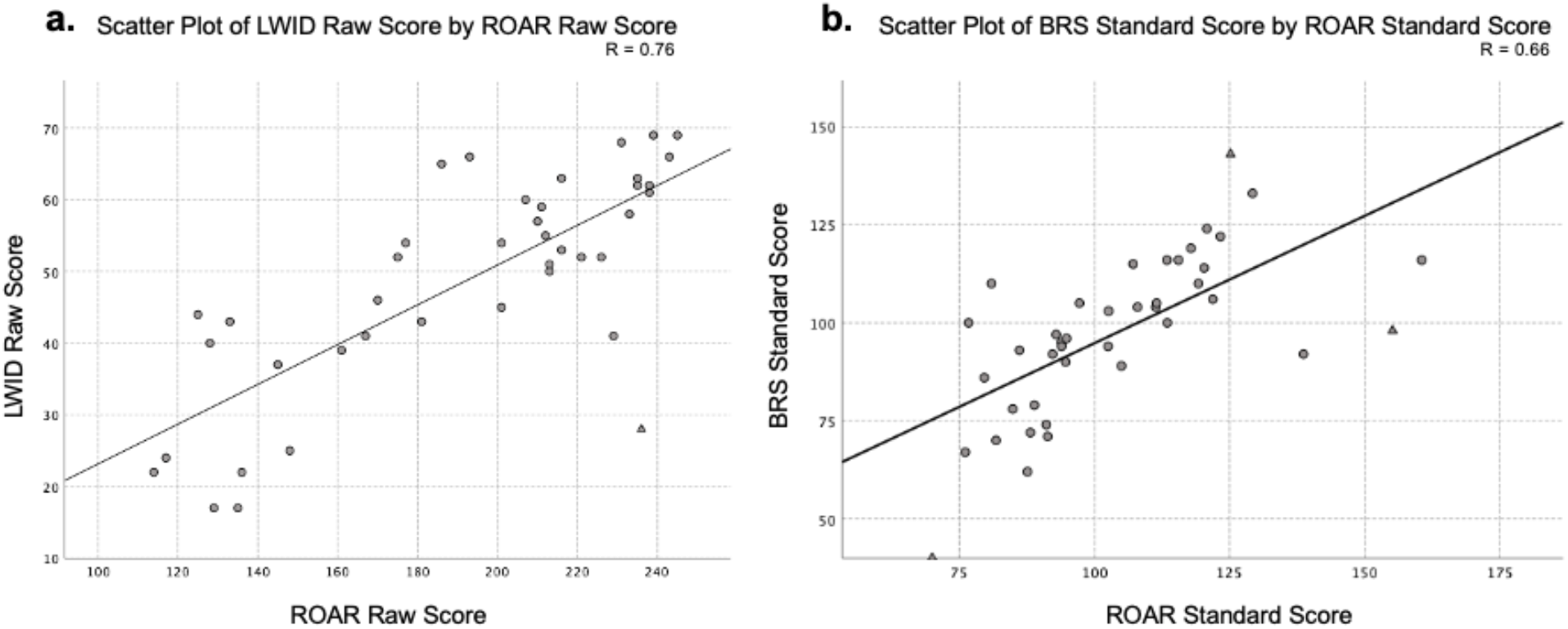
Scatter plot on the left (2a) shows each participant’s LWID raw score versus their raw score on ROAR; Pearson correlations coefficient (R = 0.76). Scatter plot on the right (2b) shows each participant’s BRS standard score versus their standard score on ROAR; Pearson correlations coefficient (R = 0.66). Triangles indicate the participants that were identified as outliers with a standardized residual that was larger than 2 (in absolute value).

### ROC curve

To evaluate ROAR for use as a screening test to classify individuals with reading difficulties, we created a receiver operating characteristic (ROC) curve and assessed area under the curve (AUC). To construct the ROC curve, we set a classification threshold for poor readers as a standard score on the BRS of less than 90. With this distinction, 26.8% (approximately the lowest quartile) of participants in our clinical sample were categorized as poor readers. The ROC curve was created by plotting true positive rate versus the false positive rate. The curve created has an area under the curve (AUC) of 0.90 (CI 0.81 to 1.00) (Figure 3). The coordinates on the ROC curve were then analyzed to determine the score on ROAR with the optimal sensitivity and specificity of distinguishing between good and poor readers. A standard score of 91.22 or greater on ROAR has a sensitivity of 90.0 and specificity of 81.8 for predicting a BRS score greater than 90.

**Figure 3:**
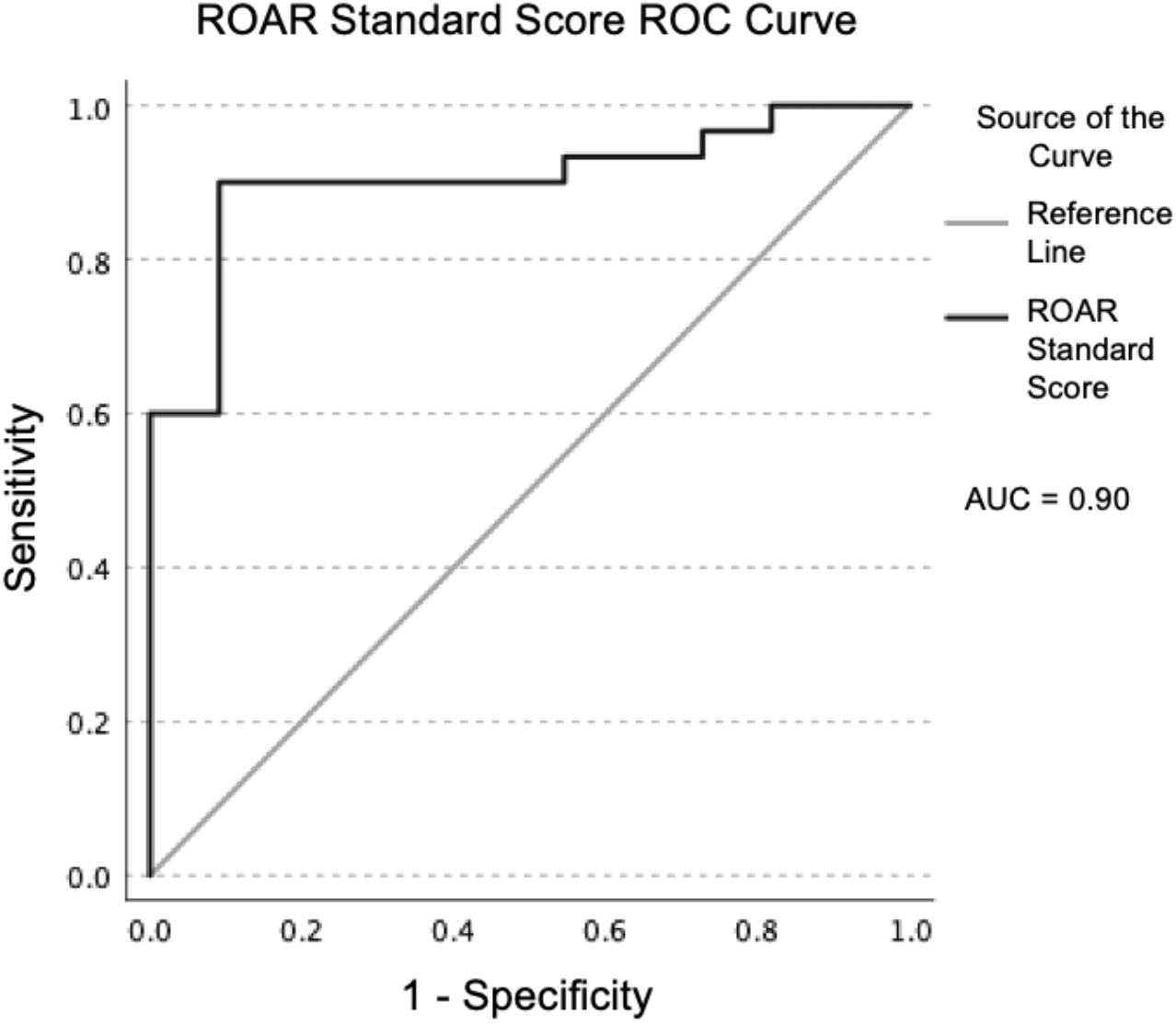
ROC curve of participants’ standard score on ROAR. Participants were classified as poor readers versus good readers based on a cutoff standard score of 90 on BRS. Area under the curve (AUC) = 0.90 (CI 0.81 to 1.00)

## DISCUSSION

In this study, we found a strong correlation between standard scores on ROAR and standard scores on the Woodcock Johnson LWID, WA and BRS. The area under the ROC curve showed that ROAR was an excellent measure to identify children at highest risk of having poor reading skills. These findings support the use of ROAR as a preclinical screening tool for identifying children who present to Developmental-Behavioral Pediatrics clinic who are at high risk for reading difficulty or reading disorder. Having an accurate screening tool to identify children at highest risk for reading disorders could be particularly helpful to DBP clinicians because reading disorders are highly prevalent in the DBP clinical population and could contribute to academic, behavioral, and social struggles in school. ROAR provides additional benefit to clinicians because it could be done prior to the clinic visit with results available to the clinician to review with the family during a comprehensive assessment.

One limitation of this study is the small sample size. Research staff had anticipated being able to recruit more participants for the study within the timeline than were ultimately recruited. The staff found that participants largely declined participation in the study because they did not want to add extra time to their clinical visit. If participants left the clinic without completing all aspects of the study, it was difficult to entice them to finish the procedures despite emailing them with reminders several times. To add more confidence in using ROAR as a screening tool in a clinical population, it should be assessed in a larger population. The benefit of a larger population would be to replicate these findings and to assess how ROAR performs as a screening tool in clinical sub-populations, which we could not accomplish confidently in the present study. In addition, the replication could use the new version of the ROAR, which uses a computer adaptive testing algorithm, reducing the time required to generate accurate results.

In this pilot study we used ROAR for children 6 years and up. However, based on our experience, we would recommend the clinical use of the ROAR for children over age 7 years because we found that younger children could not easily complete the test. This issue could also be ameliorated with further updates to the ROAR gameplay to make the task and instructions clearer and more engaging for younger participants.

Another opportunity to consider in future research with a larger sample is to investigate why discrepancies exist between ROAR score and Woodcock Johnson scores. In this clinical sample the correlation between ROAR scores and Woodcock Johnson scores was considerably lower than the correlation in the general population (r = 0.76 current study versus r = 0.91 in the general population). A larger sample would be required to determine the extent to which this reflects specific characteristics of the clinical sample versus the influence of outliers in a small sample. For example, one of the outliers had an average Woodcock Johnson score for age and an above average ROAR score. One possible explanation is that this participant’s inattention in a new clinical setting made the child perform below their true ability level when they were completing one-on-one, individually administered assessments with a clinician. In this scenario, ROAR performed in a distraction-free environment might, in fact, be a more accurate estimate of true ability than a standardized clinical measure. Another possibility is that, in certain cases, the ROAR taps into different underlying skills than the Woodcock Johnson and might, therefore, miss some cases of reading difficulties that would have been detected by an individually administered assessment. Another possibility to be aware of is that an outlier might have received help from a parent or sibling; with an unmonitored assessment, this possibility must be taken into consideration. At present, ROAR development team is making efforts to write algorithms to flag these cases. In summary, future research should pursue a large sample, evaluation of test performance in defined subgroups, use of the computer adapted testing algorithms, and detailed case studies to understand the best use cases of an automated, online screening tool in a DBP clinical setting.

## CONCLUSION

We found strong correlations between ROAR scores and a gold standard measure of reading ability and a high area under the curve in ROC analysis, providing strong preliminary support for the use of ROAR as a pre-clinical screening tool to accurately identify those children referred to DBP clinical care as likely having reading difficulties. Future research in larger samples is necessary to fully understand the strengths and weaknesses of ROAR in relation to conventional, individually administered assessments and how differences between ROAR and conventional assessment vary by clinical condition. In addition, this study provides a strong proof-of-concept for how DBP clinics can leverage new technology to improve efficiency in conducting comprehensive evaluations of young children at high risk for co-existing reading and related disorders.

## Data Availability

All data produced in the present study are available upon reasonable request to the authors

## ACKNOWLEDGEMENTS

Thank you to the children and families that participated in the study. Thank you to research staff Megumi Takada and Clementine Chou for their assistance in navigating the IRB submission and for technical support. Thank you Amy Bukhardt and Jasmine Tran for technical assistance.

## SOURCES OF SUPPORT

Wu Tsai Neurosciences Institute, Neuroscience: Translate, NIH NICHD R01HD095861 Health Resources and Services Administration T77 MC09796

Dr. Elizabeth Barrington is an Elizabeth and Russell Siegelman Postdoctoral Fellow of the Stanford Maternal and Child Health Research Institute.

